# Rationale and Design of an Artificial Intelligence Model for Diastolic Heart Failure (AID-HF): A Canadian Cardiomyopathy Collaborative (C3) Study

**DOI:** 10.64898/2026.05.27.26354226

**Authors:** Tanya Papaz, Suhani Patel, Rajadurai Akilen, Sandar Min, Robert Lesurf, Jean-Lucien Rouleau, Matthieu Ruiz, Christopher Z Lam, Andreea Dragulescu, Mark K Friedberg, Luc Mertens, Maxime Tremblay-Gravel, Andrew D Krahn, Rafik Tadros, Seema Mital

**Affiliations:** Genetics and Genome Biology Program, The Hospital for Sick Children, Toronto, Ontario, Canada; Ted Rogers Centre for Heart Research, Toronto, Ontario, Canada; Research Centre, Montreal Heart Institute, Montreal, Quebec, Canada; Department of Medicine, Faculty of Medicine, Université de Montréal, Montreal, Quebec, Canada; Department of Nutrition, Faculty of Medicine, Université de Montréal, Canada; Department of Diagnostic Imaging, The Hospital for Sick Children, Toronto, Ontario, Canada; Cardiovascular Genetics Centre, Montreal Heart Institute, Montreal, Quebec, Canada; Division of Cardiology, Department of Medicine, University of British Columbia, Vancouver, British Columbia, Canada; Labatt Family Heart Centre, Department of Pediatrics, The Hospital for Sick Children, University of Toronto, Toronto, Ontario, Canada

**Keywords:** artificial intelligence, cardiomyopathy, diastolic heart failure, proteomics, lipidomics, genomics, multiomics

## Abstract

Diastolic heart failure (HF) in primary cardiomyopathy is under-recognized and often diagnosed late, particularly in children. While recent studies have advanced understanding of HF with preserved ejection fraction in older adults, the prevalence, outcomes and molecular drivers of diastolic HF in pediatric and young adult cardiomyopathy remain poorly defined, where disease is typically driven by primary myocardial disease rather than acquired co-morbidities.

The Canadian Cardiomyopathy Collaborative (C3) was assembled to leverage three of Canada’s leading pediatric and adult cardiomyopathy biobank registries. Its flagship initiative, Artificial Intelligence to Model Diastolic Heart Failure (AID-HF), aims to integrate deep phenotyping - including comprehensive diastolic function assessment - with genomics, lipidomics and proteomics and apply machine learning to identify biological and clinical signatures that drive cardiac function and outcomes in cardiomyopathy. Harmonized phenotyping and multiomics protocols across registries will create a uniquely integrated national data resource and enable the goals of AID-HF i.e., earlier diagnosis and new therapeutic targets for diastolic HF in cardiomyopathy.

## INTRODUCTION

Cardiomyopathy and congenital heart disease are the leading causes of childhood HF with a growing population of children and young adults requiring life-long cardiac care. There are no targeted therapies, and drugs developed for adults have limited efficacy in children.^1^ Diastolic HF is characterized by impaired relaxation and/or increase ventricular chamber stiffness resulting in reduced left ventricular (LV) filling and elevated LV filling pressures.^2^ Diastolic dysfunction is often diagnosed late and under-recognized, particularly in children.^3^ Adult echocardiography guidelines often lack sensitivity and specificity^4,5^ and fail to reliably characterize diastolic dysfunction in children in part due to significant age-related changes in diastolic parameters,^2,3,6^ and differences in etiology. While adult diastolic HF is often caused by acquired conditions like ischemic heart disease, diabetes, hypertension, or obesity, diastolic HF in children is usually seen in the context of structural conditions like congenital heart disease and heritable cardiomyopathy.^7,8^

Adult studies have used machine learning to better characterize phenogroups with diastolic dysfunction by linking echocardiographic parameters, invasive hemodynamic measurements and clinical parameters.^9, 10^ However, the biological drivers of diastolic HF remain poorly understood in pediatric and adult cardiomyopathy. Previous research by us and others have identified the contribution of both rare and common genetic variants (polygenic risk) as predictors of risk and/or outcomes in cardiomyopathy, yet their relationship to diastolic HF and to outcomes has not been systematically examined.^11–15^ More recently, our multiomics analysis of myocardial tissue and plasma identified a unique myocardial transcriptomic signature associated with restrictive cardiomyopathy.^16^ Of note, diastolic dysfunction was associated with upregulation of genes involved in lipid biosynthesis and oxidation, and an abnormal peripheral lipidomic profile characterized by an elevation of saturated triacylglycerols and other lipid subclasses supporting abnormal fatty acid oxidation. A machine learning model that incorporated the transcriptomic signature improved the classification accuracy for diastolic HF compared to only clinical and imaging variables.^16^

Building on these findings, we launched the Artificial Intelligence to Model Diastolic HF (AID-HF) study in order to perform multiomic characterization of pediatric and adult cardiomyopathy by incorporating comprehensive diastolic function characterization by echocardiography, cardiac biomarkers, genome sequencing, global proteomics and lipidomics. This study will leverage a collaboration that brings together three leading multicentre Canadian cardiomyopathy registries to extend this integrated multi-omics framework across the lifespan, spanning pediatric and adult disease. The overall goal is to generate an integrated model incorporating diastolic dysfunction with clinical and biological features to predict clinical outcomes in cardiomyopathy.

## METHODS

### Canadian Cardiomyopathy Collaborative (C3)

AID-HF is a multi-centre observational cohort study utilizing biospecimens and longitudinal data available in participants across three Canadian multi-centre registries - the Heart Centre Biobank Registry (The Hospital for Sick Children, Toronto, ON)^17^, Hearts in Rhythm Organization (University of British Columbia, Vancouver, BC)^18^ and Hearts in Rhythm Organization Hypertrophic Cardiomyopathy (Montreal Heart Institute, Montreal, QC)^19^. Together, this forms the Canadian Cardiomyopathy Collaborative, under the umbrella of the Canadian Heart Function Alliance (**Figure 1**). The Heart Centre Biobank Registry (established in 2007) enrolls pediatric and adult participants with congenital heart disease and childhood onset cardiomyopathies (including at-risk individuals) from 6 Ontario hospitals. The Hearts in Rhythm Organization (established in 2015) is a national data and biobank registry enrolling from 23 specialized pediatric and adult cardiogenetic clinics across Canada. Participants targeted for enrollment include those with a known inherited heart rhythm or arrhythmogenic cardiomyopathy condition, or an unexplained cardiac arrest. The Hearts in Rhythm Organization Hypertrophic Cardiomyopathy (established in 2021) is a multi-centre registry, biobank and imaging repository enrolling participants with a clinical diagnosis of hypertrophic cardiomyopathy and/or those carrying a pathogenic/likely pathogenic gene variant predisposing to hypertrophic cardiomyopathy. All participants have consented to sharing data and samples with external researchers for ongoing and future research projects. Data and/or material transfer agreements were executed with the Hospital for Sick Children as lead site. All participating sites received approval from institutional research ethics boards, and all patients/parents/legal guardians provided informed consent (and assent where appropriate).

**Figure 1:**
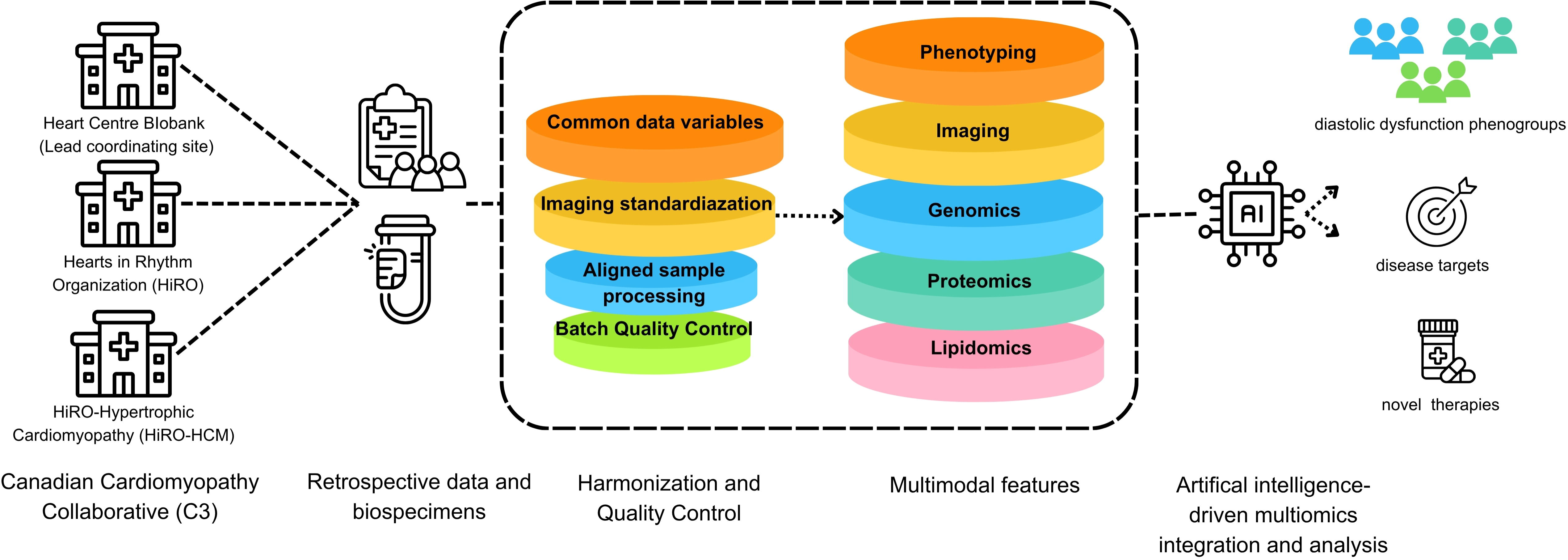
Framework of the Canadian Cardiomyopathy Collaborative and AID-HF study design.

### Study cohort for AID-HF

Inclusion criteria for AID-HF include (i) phenotype-positive participants (probands and family members) with a clinical diagnosis of primary arrhythmogenic, dilated, hypertrophic, or restrictive cardiomyopathy as further defined in **Table 1**, (ii) availability of deoxyribonucleic acid (DNA) and/or plasma and/or serum for omics assays, and (iii) informed consent for data and sample sharing. Exclusion criteria include (i) secondary causes of cardiomyopathy (i.e., syndromic, endocrine, neuromuscular, structural heart defects, inborn errors of metabolism, mitochondrial or storage disorders), (ii) other significant co-morbidities predisposing to diastolic HF i.e. diabetes, coronary artery disease and systemic hypertension requiring medical therapy, and (iii) no clinical data prior to initiation of mechanical circulatory support and/or heart transplant.

**Table 1:** Imaging criteria for diagnosis of phenotype-positive cardiomyopathy.

| **Cardiomyopathy type** | **Supporting imaging criteria (Echocardiography +/- Cardiac MRI)** |
| --- | --- |
| Arrhythmogenic cardiomyopathy (ACM) | Clinical and/or pathological diagnosis (RV-dominant, LV-dominant or biventricular based on task force criteria)<sup>33</sup>. |
| Dilated cardiomyopathy (DCM) | LV dilation and/or dysfunction defined as LVEDD z-score of $\geq 2.5$ (or $>2$ at two consecutive measurements) and/or LVEF $<50\%$ (or $<55\%$ at 2 consecutive measurements) in the context of a clinical diagnosis of DCM. |
| Hypertrophic cardiomyopathy (HCM) | Abnormal LV hypertrophy not explained by loading conditions i.e. IVSd or LVPWd z score of $\geq 2.5$ (or $>2$ at two consecutive measurements) in the context of a clinical diagnosis of HCM. For adult-sized teenagers and adults, max LV wall thickness at end-diastole $>15$ mm (or between 13-15mm in at risk individuals). |
| Restrictive cardiomyopathy (RCM) | Atrial dilation (atrial end-systolic diameter z-score $>2$) and/or abnormal LV diastolic function (without significant LV dilation or LV hypertrophy) in the context of a clinical diagnosis of RCM. |

*IVSd, interventricular septal diameter; LV, left ventricular; LVEDD, left ventricular end-diastolic diameter; LVEF, left ventricular ejection fraction; LVPWd, left ventricular posterior wall diameter; RV, right ventricular*

### Clinical Data

#### Phenotyping

Clinical data variables selected for study are based on the study objectives and their availability across participating registries and will be determined through a systematic review of data dictionaries and variable inventories. Variables that are consistently defined across registries and essential for characterizing the study population, exposures, and outcomes of interest will be prioritized. Where differences in variable definitions or collection methods exist, variables will be assessed for feasibility of harmonization prior to inclusion. This approach ensures alignment of data selection with study objectives while maximizing data completeness, comparability, and analytic validity across registries. The prioritized clinical data variables are illustrated in **Figure 2** and include demographics, cardiomyopathy type, family history, genetic etiology, symptoms, medications, laboratory markers (N-terminal pro-B-type natriuretic peptide, creatinine), and cardiac assessment (electrocardiogram, ambulatory electrocardiogram monitor, echocardiogram, exercise stress test, cardiac magnetic resonance imaging). HF related clinical outcomes include death, cardiac transplant, mechanical circulatory support, and other major adverse cardiac events including interventions like implantable cardioverter defibrillator or pacemaker. Since the primary outcome is transplant-free survival, participant clinical data from date of first clinical encounter or evaluation at a participating site until last reported evaluation or censoring event of death, transplant, or mechanical circulatory support will be utilized. Detailed data variables are listed in **Table 2**.

**Figure 2:**
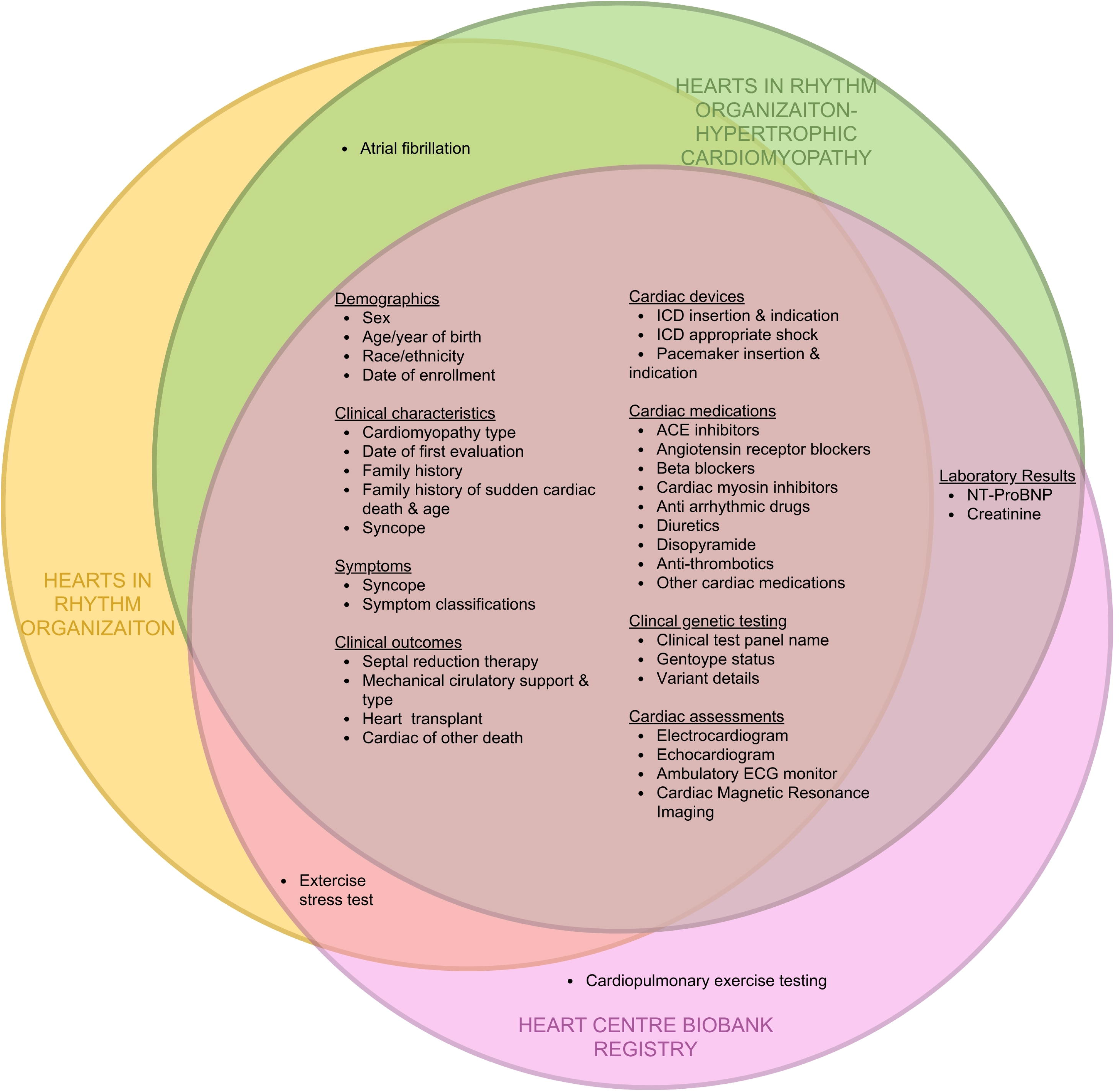
Clinical data variables across C3 registries for the AID-HF Study. ACE, angiotensin-converting enzyme; ECG, electrocardiogram; ICD, implantable cardioverter defibrillator; NT-proBNP, N-terminal pro-brain natriuretic peptide

**Table 2:** Clinical data variables and research measures.

|  | Clinical | Research |
| --- | --- | --- |
| **Demographics** |  |  |
| Sex, year of birth, race/ethnicity, date of registry enrollment | X |  |
| **Clinical characteristics** |  |  |
| Cardiomyopathy type | X |  |
| Date of first clinical encounter or evaluation | X |  |
| Family history of cardiomyopathy in first degree relative | X |  |
| Family history of sudden cardiac death in first degree relative & age | X |  |
| **Symptoms with dates** |  |  |
| History of unexplained syncope | X |  |
| Symptom classifications | X |  |
| **Clinical outcomes with dates** |  |  |
| Septal reduction therapy (myectomy, alcohol septal ablation) | X |  |
| Mechanical circulatory support with type (ventricular assist device, extracorporeal membrane oxygenation) | X |  |
| Heart transplant | X |  |
| Cardiac or other death | X |  |
| **Cardiac devices with dates** |  |  |
| ICD insertion & indication | X |  |
| Appropriate ICD discharge/shock | X |  |
| Pacemaker insertion & indication | X |  |
| Cardiac resynchronization therapy | X |  |
| **Medications** |  |  |
| Angiotensin-converting enzyme inhibitors, Angiotensin receptor blockers, Angiotensin receptor-neprilysin inhibitors, Beta blockers, Cardiac myosin inhibitors, Anti-arrhythmic drugs, Diuretics, Disopyramide, Anti-thrombotics, other cardiac medications | X |  |
| **Laboratory results with dates** |  |  |
| N-terminal pro-brain natriuretic peptide | X | X |
| Creatinine | X | X |
| **Clinical genetic testing with dates** |  |  |
| Clinical genetic test panel name | X |  |
| Genotype status (pathogenic/likely pathogenic, variant of uncertain significance, none/benign) | X |  |
| Variant details (affected gene, pathogenicity, nucleotide substitution, amino acid change, mutation type/impact, zygosity, inheritance) | X |  |
| **Cardiac assessment with dates** |  |  |
| Electrocardiogram (heart rate, PR interval, QRSd, QTc) | X |  |
| Echocardiogram (height, weight, systolic and diastolic blood pressure. Measures listed in Table 3) | X | X |
| Ambulatory ECG monitor (atrial fibrillation, non-sustained ventricular tachycardia, ventricular tachycardia) | X |  |
| Exercise stress test (type of test, presence of abnormal response to exercise including blunted BP, blunted HR, ectopy, ischemia) | X |  |
| Cardiopulmonary exercise test (heart rate, blood pressure, respiratory exchange ratio, peak workload & percent predicted for normal, peak $\text{VO}_2$, $\text{VO}_2$ at anaerobic threshold & percent predicted for normal values, $\text{V}_E/\text{VCO}_2$ slope) | X | |
| Cardiac magnetic resonance imaging (height, weight. Measures listed in Table 3) | X | X |
| **Biological data** |  |  |
| Whole genome sequencing |  | X |
| Proteomics |  | X |
| Lipidomics |  | X |

*ECG, electrocardiogram; ICD, implantable cardioverter defibrillator; PR, P wave to R wave; QRSd, QRS complex duration; QTc, corrected QT interval; $\text{VO}_2$ , volume of oxygen; $\text{V}_E/\text{VCO}_2$ , minute ventilation/carbon dioxide production*

### Imaging

#### Echocardiography

Clinically acquired echocardiographic studies captured using standardized clinical protocols are available from registry participants and will be analyzed at research echocardiographic core labs (The Hospital for Sick Children and Montreal Heart Institute) through overreading of echocardiographic data stored in Digital Imaging and Communications in Medicine format. The core labs will measure systolic and diastolic function parameters and chamber sizes (**Table 3**). Resting transthoracic echocardiograms in sinus rhythm will be selected for evaluation; transesophageal or exercise echocardiograms will be excluded.

**Table 3:**
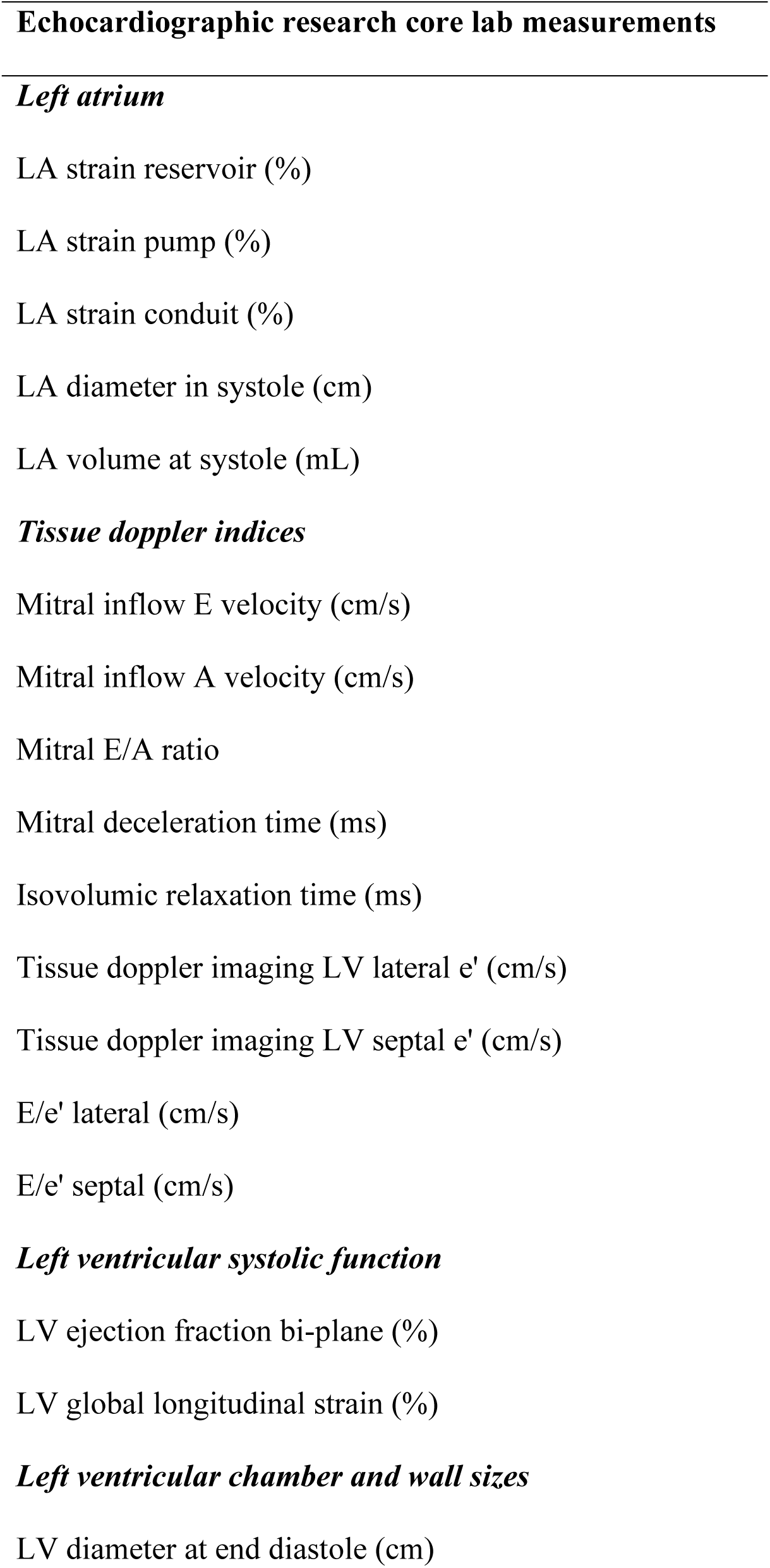
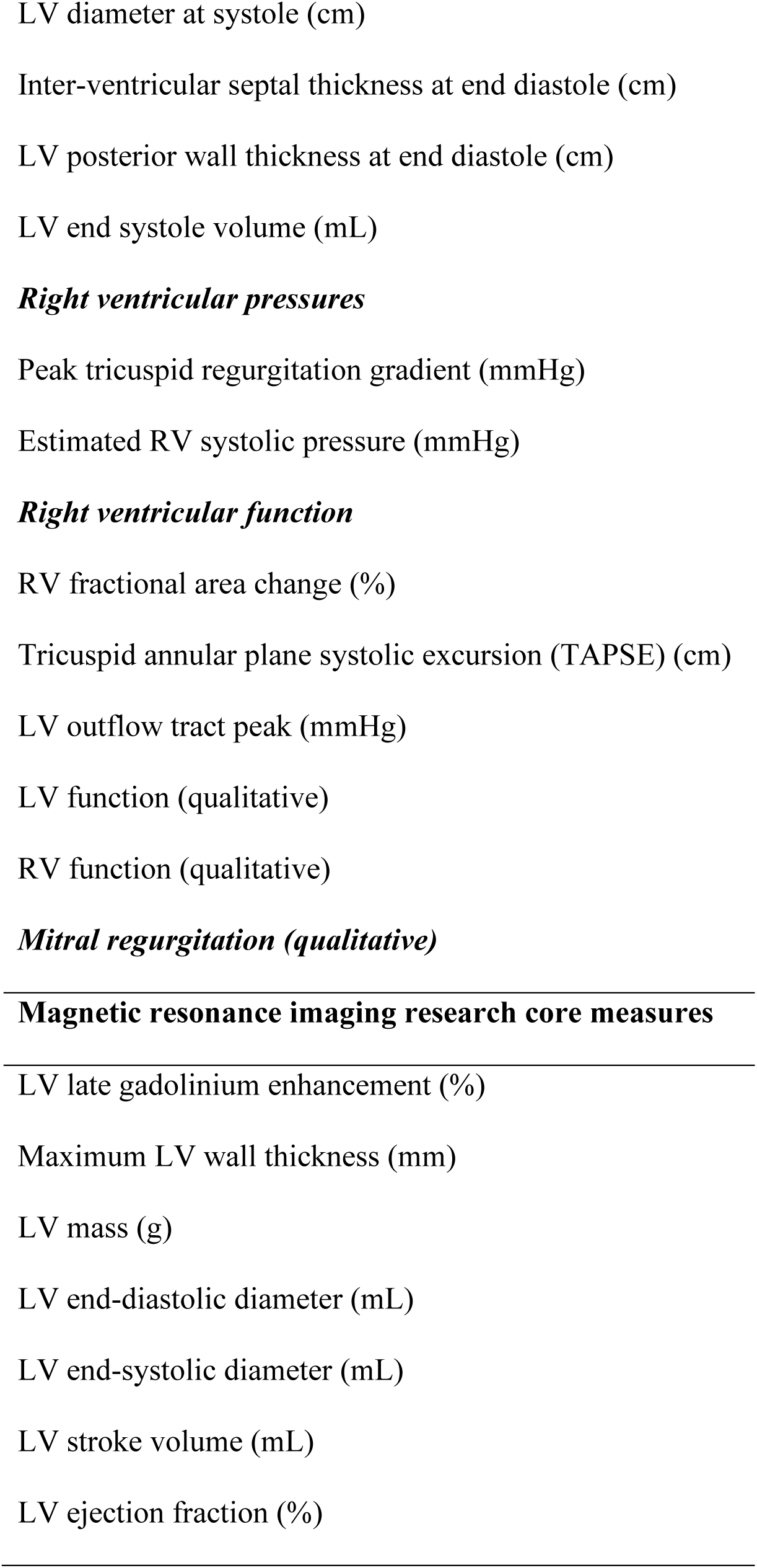
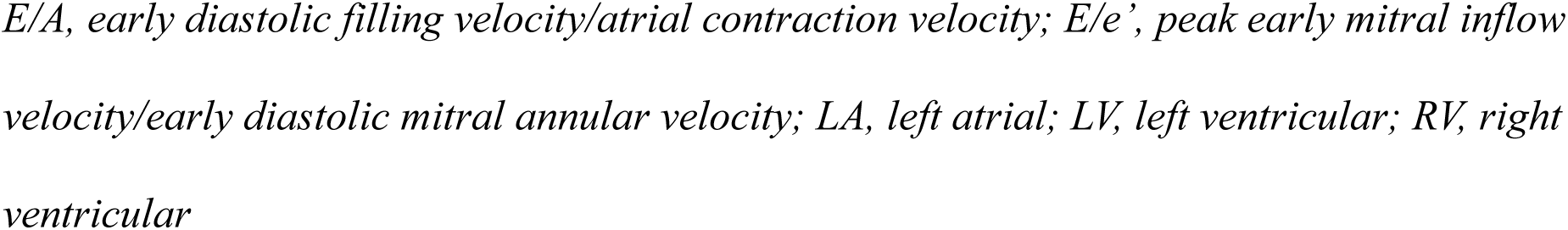
Research imaging core lab measurements.

Echocardiographic images will be analyzed using software for offline analysis (TomTec Imaging Systems, GmbH, Unterschleißheim, Germany; and EchoPAC, GE Vingmed Ultrasound AS, Horten, Norway).^20,21^ Echocardiographic parameters will be normalized using Pediatric Heart Network and/or Boston z-scores in pediatric patients and indexed to body surface area in adults.^22,23^ Where echocardiographic images are not available, only clinically reported echocardiographic measures will be used.

#### Cardiac Magnetic Resonance Imaging (CMR)

LV late gadolinium enhancement (LGE) and LV volumetric measures will be collected from CMR clinical reports from each registry, and re-evaluated using images stored in Digital Imaging and Communications in Medicine format at The Hospital for Sick Children CMR Research Core. Several LGE assessment techniques and quantification methods will be employed and evaluated for fit and reproducibility. CMR images will be analyzed and interpreted using dedicated post-processing software Medis Suite MR (Medis Medical Imaging Systems B.V., Leiden, The Netherlands).^24^ **Table 3** lists the parameters prioritized for assessment. Where applicable, values indexed to body surface area will be calculated.

### Biospecimens

Sample collection protocols across the registries will be compared to optimize joint processing (**Table 4**). For whole genome sequencing, DNA is derived from blood or saliva and collected at time of registry enrollment. Blood for DNA extraction is collected in dipotassium ethylenediaminetetraacetic acid (K_2_ EDTA) BD Vacutainer® tubes (Becton, Dickinson and Company, Franklin Lakes, NJ, USA) while saliva is collected in DNA Genotek Oragene collection kits (DNA Genotek Inc., Ottawa, ON, Canada). Blood is either sent for immediate DNA extraction or stored at -70°C in polypropylene tubes prior to batch DNA extraction. Extracted DNA is stored at -20°C with aliquots stored at 4°C. Plasma and/or serum samples are collected at registry enrollment in adults and prior to surgical intervention or clinical blood draw in pediatric patients. Plasma is collected in K_2_ EDTA vacutainers or in lithium heparin plasma separator vacutainers. Serum is collected in clot activator (silicone coated) serum vacutainers. Samples are generally processed within 30 minutes of collection and centrifuged at a force ranging from 1300-1900g for 15 minutes at room temperature. Plasma and serum are stored at - 70°C degrees or lower. There is minimal variability in centrifugation protocols across registries.

**Table 4:**
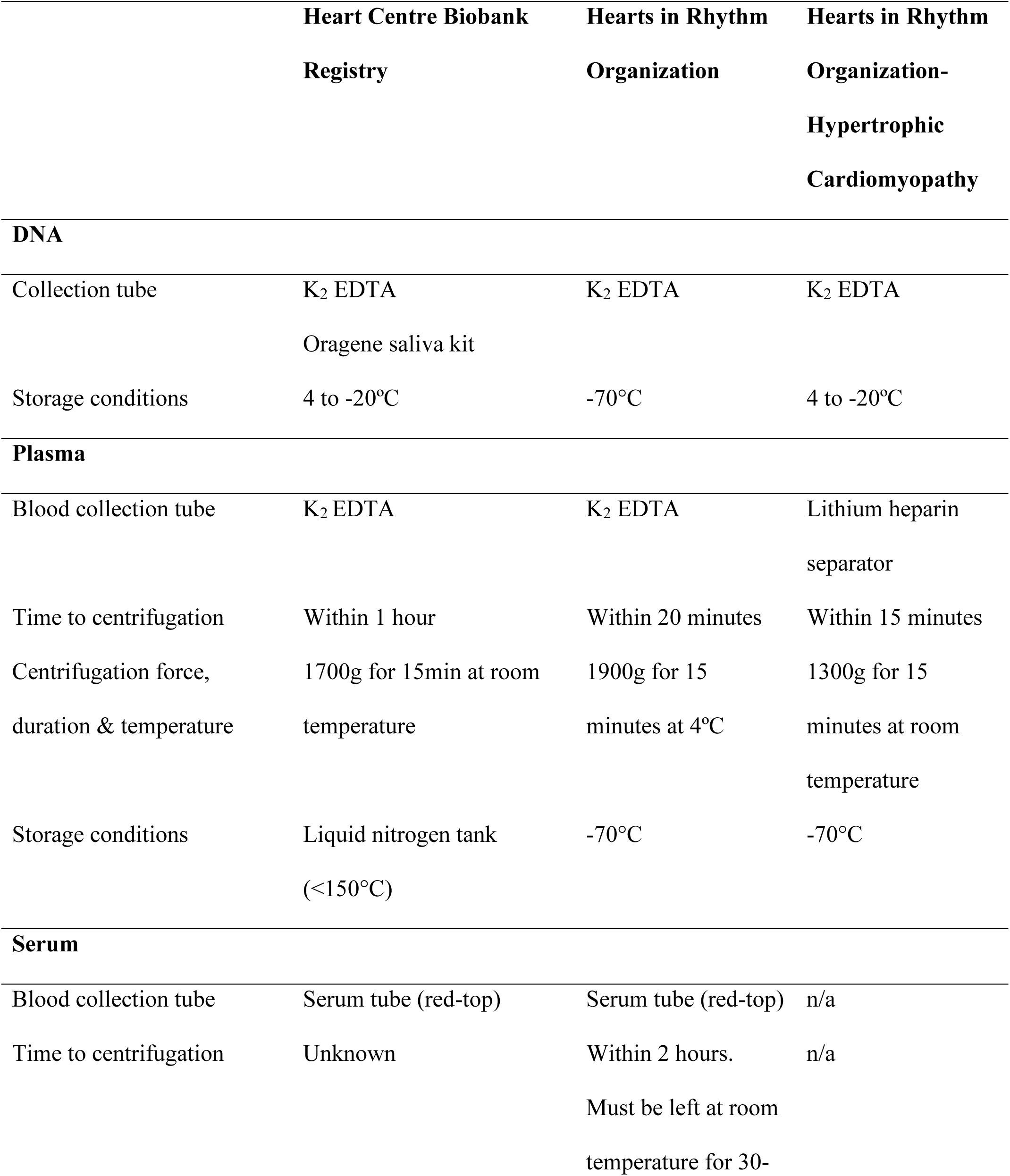
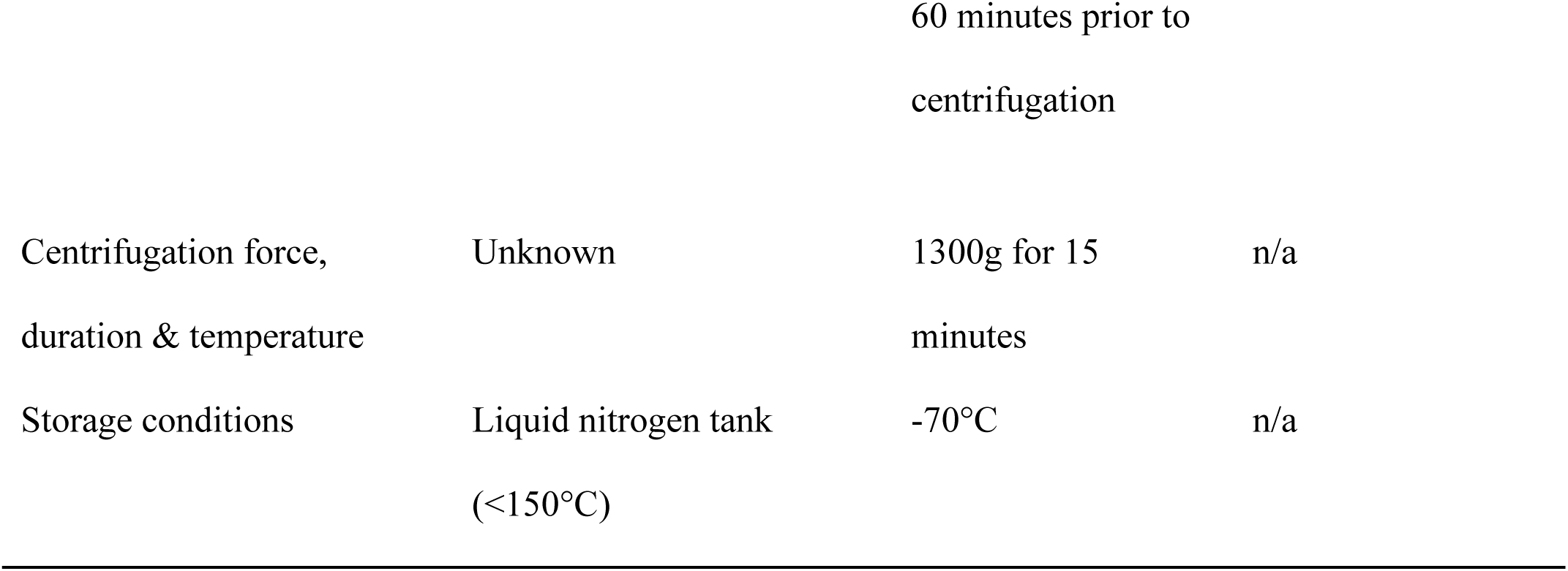
Registry sample collection protocols.

#### Genomics

DNA will be sent for whole genome sequencing to The Centre for Applied Genomics at The Hospital for Sick Children (Toronto, Ontario, Canada). Polymerase chain reaction-free library preparation and quality control will be performed prior to sequencing. Samples meeting quality control thresholds will be sequenced on the Illumina NovaSeq X Series platform producing an average autosomal depth of 30X coverage (Illumina, Inc., San Diego, CA, USA).^25^ Alignment and variant calling pipelines will be harmonized. Quality control of genome sequences including variant calling, prioritization and interpretation will be performed in line with methods previously published.^26,27^

#### Proteomics

Proteins from plasma or serum will be assayed at the High-Throughput Screening facility at the Lunenfeld Tanenbaum Research Institute at Mount Sinai Hospital (Toronto, Ontario, Canada) using the Olink® Reveal Assay (Olink Proteomics, Uppsala, Sweden) which measures 1,034 actionable and druggable proteins detectable in biofluids.^28^ The panel is enriched for cardiometabolic, inflammation, immune-related and other pathways. Olink Reveal uses Proximity Extension Assay™ technology coupled to next-generation sequencing. For each protein, two antibodies labeled with unique DNA oligonucleotides bind simultaneously to the target, bringing the oligos into proximity. The oligos hybridize, are extended, and amplified by polymerase chain reaction to generate a protein-specific DNA barcode. Each amplicon also contains protein-specific barcodes, sample indices, and sequencing adaptors. Samples across all plates will be randomized by registry, diagnosis, sample type and age and prepped in accordance with Olink Reveal lab instructions.^28^ All samples meeting quality control thresholds by bioanalyzer will be sequenced on the Illumina NovaSeq X Series platform.

#### Lipidomics

Lipids from plasma or serum samples will be processed at the Montreal Heart Institute (Montreal, Quebec, Canada) as previously described.^29–31^ Lipids will be extracted into a methyl tert-butyl ether mixture, followed by extraction, drying and solubilization. The samples will be diluted before injection into a 1290 Infinity High-Performance Liquid Chromatography coupled with a 6530 accurate mass Quadrupole Time-of-Flight mass spectrometry system (Agilent Technologies Inc., Santa Clara, CA, USA) via a dual electrospray ionization source in positive scan mode. Prior to extractions for this study, a pilot was performed that confirmed no significant difference in the lipidomic profile when comparing blood collected in K_2_ EDTA (Heart Centre Biobank Registry; Hearts in Rhythm Organization) versus lithium heparin (Hearts in Rhythm Organization-Hypertrophic Cardiomyopathy). Extractions will be performed consecutively and independently for samples collected from each registry accounting for differences in collections. Batch corrections will be applied when datasets are jointly analyzed.

### Analysis Plan

#### Data preprocessing

Feature input will include biological data (variant burden of rare pathogenic or likely pathogenic cardiomyopathy-associated variants as well as common variants i.e., polygenic risk score associated with LV function, log-transformed and normalized expression values of genes, proteins, metabolites) and clinical variables including cardiac imaging and laboratory test results. Continuous variables will be standardized. Categorical variables will be encoded as binary indicator variables. Missing data will be handled using multiple imputations where appropriate.

#### Supervised Machine Learning

Data pooled from all sites will be spilt into training and testing cohorts using a 70:30 split, with stratification to preserve outcome distribution. Machine-learning approaches will be applied to identify clinical and biological markers associated with clinical outcomes. These will include individual diastolic function parameters corrected for changes related to growth. Clinical outcomes, including a composite of mechanical circulatory support, heart transplantation, and all-cause mortality, will be included as primary endpoints to assess the prognostic relevance of identified markers. Model hyperparameters will be optimized using randomized search with k-fold cross-validation. Feature importance will be assessed using Shapley Additive Explanations values. Model performance in predicting outcomes will be evaluated in the training and testing cohorts. Model discrimination will be assessed using the Area Under the Receiver Operating Characteristic curve and Area Under the Precision–Recall Curve. Patient phenogroups will be incorporated into the analysis where there is evidence of confounding biological differences that contribute to outcome. Top-ranked clinical, imaging and omics markers with strong discriminating accuracy for primary outcome will be prioritized for downstream analyses.

#### Unsupervised Machine Learning

Unsupervised approaches will be used to identify phenogroups of shared clinical and biological features. Patient similarity matrices will be generated for each data modality using a normalized distance metric and transformed into affinity networks. These modality-specific networks will be integrated using Similarity Network fusion techniques, generating a unified patient similarity network.^32^ Clustering methods will be applied to the fused network to identify phenogroups, with the optimal number of clusters determined based on stability metrics. After phenogroups have been generated and clustered, clinical outcomes will be compared between phenogroups.

This approach will enable identification of clinical, imaging and molecular markers that are associated with adverse clinical outcomes. Secondary analyses will evaluate differential responses of identified phenogroups to heart failure therapies, supporting future development of phenotype-guided strategies for cardiomyopathy.

## DISCUSSION

The C3 brings together Canadian pediatric and adult cardiomyopathy registry participants to allow investigation and better understanding of diastolic HF across the lifespan. Cardiomyopathy reflects primary muscle disease that is often genetic, thereby allowing an opportunity to understand the biological underpinnings of diastolic dysfunction without the confounding effect of aging and comorbidities seen in adult HF with preserved ejection fraction. By harmonizing deep clinical phenotyping with genomics, proteomics, and lipidomics at scale, AID-HF addresses a critical gap in defining the unique clinical and biological signatures that drive cardiac dysfunction and outcomes in cardiomyopathy across all ages. Jointly studying children and adults with primary cardiomyopathy without significant co-morbidities will allow identification of diastolic abnormalities and separation of developmental and degenerative processes while limiting survivor bias. The inclusion of various cardiomyopathy subtypes will also enhance biologically informed phenogrouping that integrates structure, mechanism and physiology.

Through combining pre-existing data assets in biobank registries, the C3 systematically harmonizes heterogeneous datasets through a structured data alignment process designed to enable cross-registry comparability. Variable definitions for common data variables will be mapped to common data standards using harmonized metadata frameworks where quality control procedures are applied to resolve discrepancies and ensure consistency. Because methods for defining and grading diastolic HF differ between pediatric and adult populations and are not well established in pediatrics, individual diastolic function parameters will be incorporated into analytical models rather than relying on categorical diastolic dysfunction grading. Variability in sample collection time points and collection methods across registries will be explicitly considered during sample processing, with protocols adapted where such differences are expected to influence downstream measurements. In addition, batch correction will be applied to processed datasets where needed to enable meaningful comparison across samples while minimizing non-biological sources of variation. The use of central core labs for imaging assessment as well as for sample processing and omics data generation will further enable integration of data across registries. This approach enables the integration of heterogenous datasets into a unified analytical resource but also provides a scalable template for future efforts seeking to link independent registries while maintaining data integrity, interoperability, and reusability across research initiatives.

Registry data including core lab generated imaging data and biological data will be available to the scientific community, with access managed through existing registry governance structures, oversight committees, and data-sharing policies of each contributing registry. A governance framework will be developed to enable coordinated stewardship and access to data from the three different registries. A key objective at study completion is to provide access via a federated data access platform through which data assets can be viewed and accessed collectively. Such a model would enable cross-registry visibility and analysis while preserving local control, regulatory compliance, and registry-specific governance requirements.

A major strength of the study is its ability to integrate and standardize data across diverse national registry platforms, creating a uniquely powerful resource to identify shared clinical and biological signatures across age groups. This lifespan precision medicine approach involving artificial intelligence and integrated multiomics has the potential to transform the field by enabling earlier diagnosis, improved risk stratification, and the discovery of novel therapeutic targets for cardiomyopathy.

## Data Availability

All data produced in the present study are available upon reasonable request to the authors

## ACKNOWLEDGEMENTS

We are indebted to the tireless work of the Hearts in Rhythm Organization, Hearts in Rhythm Organization Hypertrophic Cardiomyopathy and Heart Centre Biobank Registry study coordinators and to our patients/guardians who gladly participate to advance our understanding of heart disease, cardiac arrest and inherited arrhythmias (Clinical trial registration: NCT04189822; NCT05100420; theheartcentrebiobank.com).

## PATIENT CONSENT STATEMENT

### Ethics Statement

The study adheres to the relevant ethical guidelines. Research ethics board approval was obtained at each participating institution - The Hospital for Sick Children (REB# 1000013735), Montreal Heart Institute (REB# 2025-3479) and the University of British Columbia (REB# H24-00958).

### Patient Consent

All three registries obtained informed consent and assent from patient and/or parent/legal guardian.

### Declaration of generative AI and AI-assisted technologies in the manuscript preparation process

During the preparation of this work the author(s) used a large language model (M365 Copilot) on occasion to assist with language editing and rephrasing. After using this tool/service, the author(s) reviewed and edited the content and take(s) full responsibility for the content of the published article.

## ABBREVIATIONS

AID-HF: Artificial Intelligence Model for Diastolic Heart Failure
C3: Canadian Cardiomyopathy Collaborative
CMR: Cardiac Magnetic Resonance Imaging
DNA: deoxyribonucleic acid
HF: Heart failure
K_2_ EDTA: dipotassium ethylenediaminetetraacetic acid
LGE: Late gadolinium enhancement
LV: Left ventricular
REB: Research ethics board

